# Endovascular treatment of ruptured very small intracranial aneurysms: a systematic review and meta-analysis

**DOI:** 10.1101/2023.05.16.23290074

**Authors:** Hidetoshi Matsukawa, Sameh Samir Elawady, Mohammad Mahdi Sowlat, Sami Al Kasab, Kazutaka Uchida, Shinichi Yoshimura, Alejandro M Spiotta

## Abstract

**Background:** Endovascular treatment (EVT) is widely accepted for intracranial aneurysms due to its safety and efficacy. However, EVT of ruptured very small intracranial aneurysms (RVSIA) (≤3 mm) is still challenging and the risk-benefit ratio of EVT remains unclear. The aim of this study was to evaluate the safety and efficacy of EVT of RVSIA.

**Methods:** We performed a systematic review and meta-analysis of the studies on EVT of RVSIA. Pooled prevalence rates were calculated for initial and follow-up complete occlusion rates (Raymond Roy Grade 1), recanalization, retreatment, long-term favorable outcome (modified Rankins scale score 0 to 2 or Glasgow Outcome Scale 4 or 5), procedure-related complications (coil herniation, thromboembolism, and intraprocedural re-rupture), and procedure-related mortality. Pooled odds ratios were calculated to compare these outcomes between simple coiling and stent-assisted coiling (SAC).

**Results:** Of the 600 studies screened, 24 studies with a total of 1355 RVSIAs treated with EVT were included. The initial and follow-up complete aneurysm occlusion rates were 64% (95% confidence interval [CI]: 52–74%) and 85% (95% CI: 74–92%). The rates of recanalization and retreatment were 6% (95% CI: 3–10%) and 3% (95% CI: 2–4%). The favorable long-term follow-up outcome was observed in 91% (95% CI: 89– 93%) of patients. The rates of coil herniation, thromboembolism, and intraprocedural rupture were 2% (95% CI: 1–8%), 4% (95% CI: 3–6%), and 4% (95% CI: 2–7%), respectively. Mortality was 3% (95% CI: 2–4%). Comparison of outcomes between simple coiling and SAC revealed no significant difference, except for a higher likelihood of recanalization in the coiling group (Odds ratio, 3.51 [95% CI, 1.31–9.45]).

**Conclusions:** Our meta-analysis demonstrates that EVT for RVSIA is a feasible, effective, and safe approach that is associated with favorable clinical outcomes in both the short and long term.

## Introduction

The incidence of aneurysmal subarachnoid hemorrhage (aSAH) is approximately 6–11 per 100 000 persons per year.^1, 2^ In spite of the understanding of the pathophysiology, around 30% of patients who survive after aSAH will not regain full independence.^3^ Very small intracranial aneurysm (≤ 3 mm) represents 13.2% - 15.1% of all intracranial aneurysms.^4, 5^ Although the risk of rupture is known to increase with the increasing size of an aneurysm^6, 7^, a large number of patients have subarachnoid hemorrhage as a result of rupture of intracranial aneurysms with diameters < 5 mm.^8–12^ Very small aneurysms may be too small to accept a clip without narrowing or tearing the parent vessel.^13, 14^ Endovascular treatment (EVT) of such aneurysms is also considered to be challenging due to the thin fragile aneurysm wall, with limited space to obtain a stable microcatheter position for coil deployment, and with a high risk of intraprocedural rupture.^4, 15–18^ However, recent advances in endovascular techniques and devices have led to the rapid evolution of coil embolization techniques for intracranial aneurysms, which might allow for safer coil approach to ruptured very small intracranial aneurysm (RVSIA). In fact, some studies demonstrated successful EVT of RVSIA with stent-assisted coiling (SAC)^19–21^ although the number of patients was small to reach a meaningful conclusion. Therefore, the risk-benefit ratio of EVT of RVSIA remains unclear. To examine the contemporary safety and efficacy of EVT of RVSIA and to compare outcomes between primary coiling and SAC, we aimed to systematically review the literature and conduct a meta-analysis.

## Material and Methods

### Search strategy

This study was conducted according to the Preferred Reporting Items for Systematic Reviews and Meta-Analyses (PRISMA) guidelines. A comprehensive search of PubMed and Scorpus databases up to February 18, 2023, was conducted to identify relevant studies. We used the following keywords: “burst,” “ruptured,” “small,” “tiny,” “3mm,” “aneurysm,” “brain,” “intracranial,” “berry,” “cerebral,” “circle of Willis,” “saccular,” “fusiform,” “embolize*,” “coil,” “stent,” and “endovascular.” These keywords were combined using “OR” or “AND.”

### Selection criteria

Our study included original articles that reported the outcomes of EVT for ruptured very small intracranial aneurysms without any restrictions on age, sex, race, ethnicity, country of origin, or publication date. The article was not eligible if it met any of the following exclusion criteria: (1) aneurysm size > 3mm; (2) blood blister and dissecting aneurysms; (3) unreliable, non-extractable, duplicated, and overlapped data sets; (4) no full text available; (5) conference/poster abstracts; (6) case reports/series of ≤ 10 cases; or (7) review articles. Studies were identified based on the abovementioned eligibility criteria by two independent reviewers. Any disagreement was resolved through cooperative discussion.

### Data extractions

Two independent reviewers evaluated the studies and extracted the relevant the data. The extracted data included the first author, year of publication, study design, number of patients with coiling or SAC, age, gender, posterior circulation aneurysm, Hunt-Hess grade^22^, size of sac and neck, and angiographic follow-up period (month), technical success, initial and long-term complete occlusion rates, recanalization rate, retreatment rate, long-term favorable neurologic outcome, intraoperative complications (intraprocedural parent artery vasospasm, coil herniation, thromboembolic complications, and intraprocedural re-rupture), and mortality. Any discrepancies between reviewers were resolved through discussion. Complete aneurysm occlusion was defined as Raymond-Roy occlusion classification I.^23^ Favorable outcome was set as modified Rankin scale 0 to 2^24^ or Glasgow Outcome Scale 4 or 5.^25^

### Quality assessment of the included studies

Risk of bias in each included study was assessed by two independent reviewers using the Risk of Bias In Non-randomized Studies of Interventions (ROBINS-Ⅰ) tool. This tool evaluates the methodological quality of the studies in various domains, including confounding factors, selection of participants into the study, classification of interventions, deviations from intended interventions, missing data, measurement of outcomes, and selection of the reported results.^26^

### Statistical analysis

We analyzed all the data using R software version 4.2.3. A “meta” package was used to calculate pooled prevalence (%) or pooled odds ratios, and their corresponding 95% confidence intervals (CI) using either a fixed-effect or random-effect model, depending on the degree of heterogeneity present among the included studies. Heterogeneity was evaluated using Q statistics and I2 test. We considered I2 value > 50% or P-value < 0.05 to indicate substantial heterogeneity. Due to the non-comparative nature of most included studies and the small number of comparative studies (< 10 per analysis), we were unable to test for publication bias or perform meta-regression.

## Results

### Literature Search

600 studies were initially identified, 470 were excluded after title and abstract screening. 130 studies were assessed for full-text eligibility, and 110 were excluded. Additional 4 studies were identified through manual search of the references of included studies. 24 studies were included in the final analysis. A study flow diagram is shown in Fig 1.

**Figure 1.**
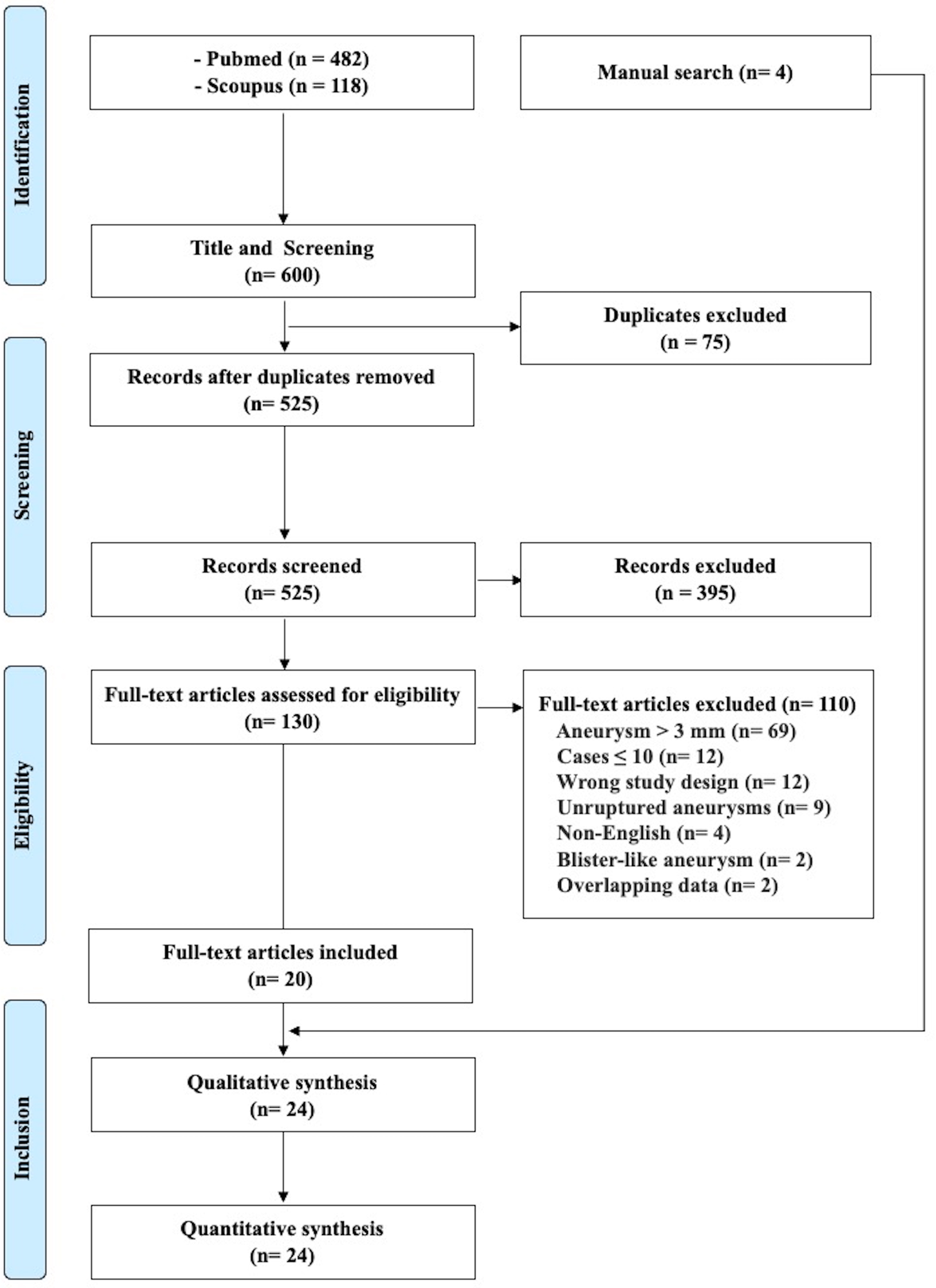
Flow diagram of the search strategy and study selection

**Figure 2.**
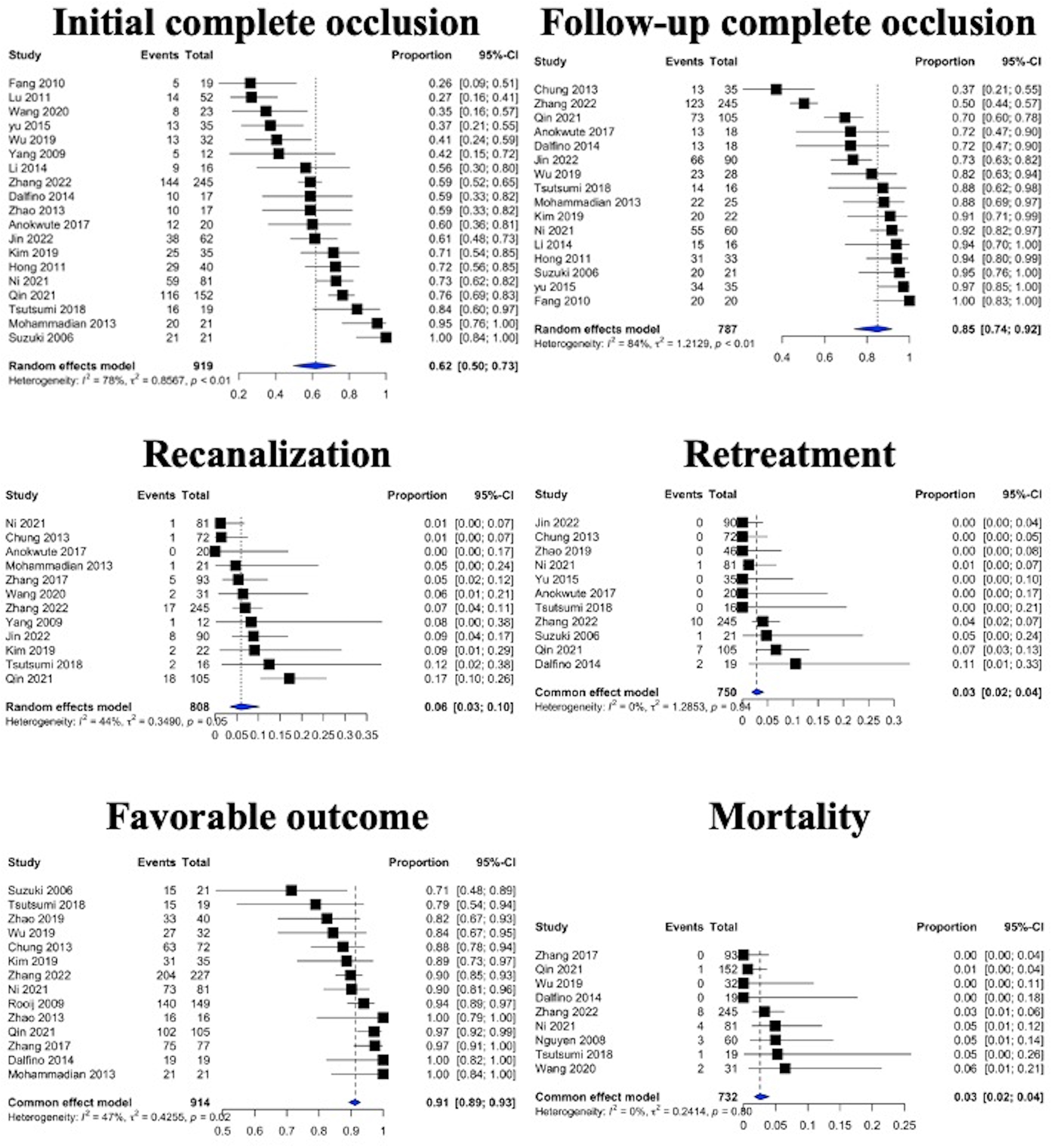
The pooled prevalence of radiological and clinical outcomes

**Figure 3.**
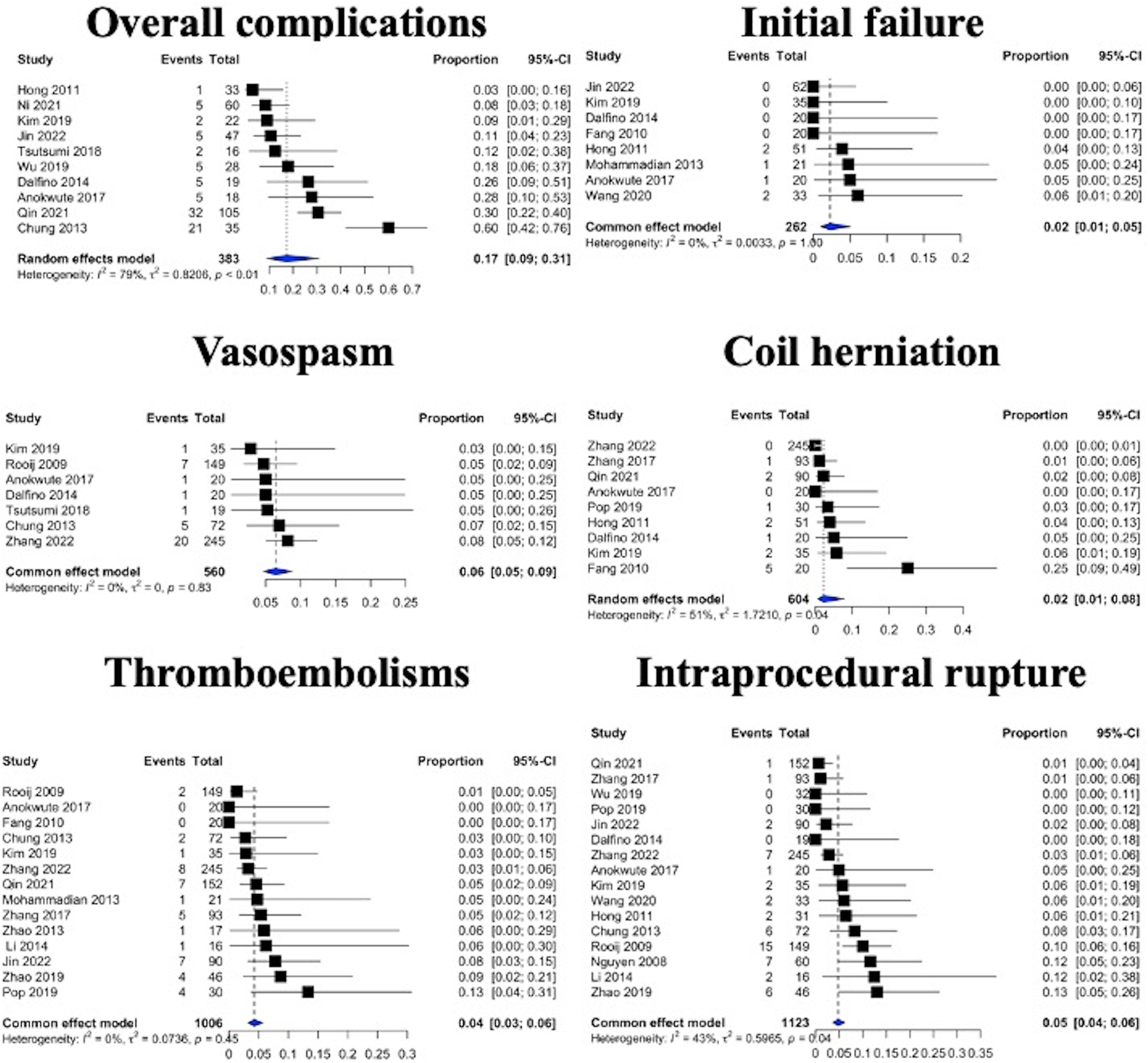
The pooled prevalence of adverse events

**Figure 4.**
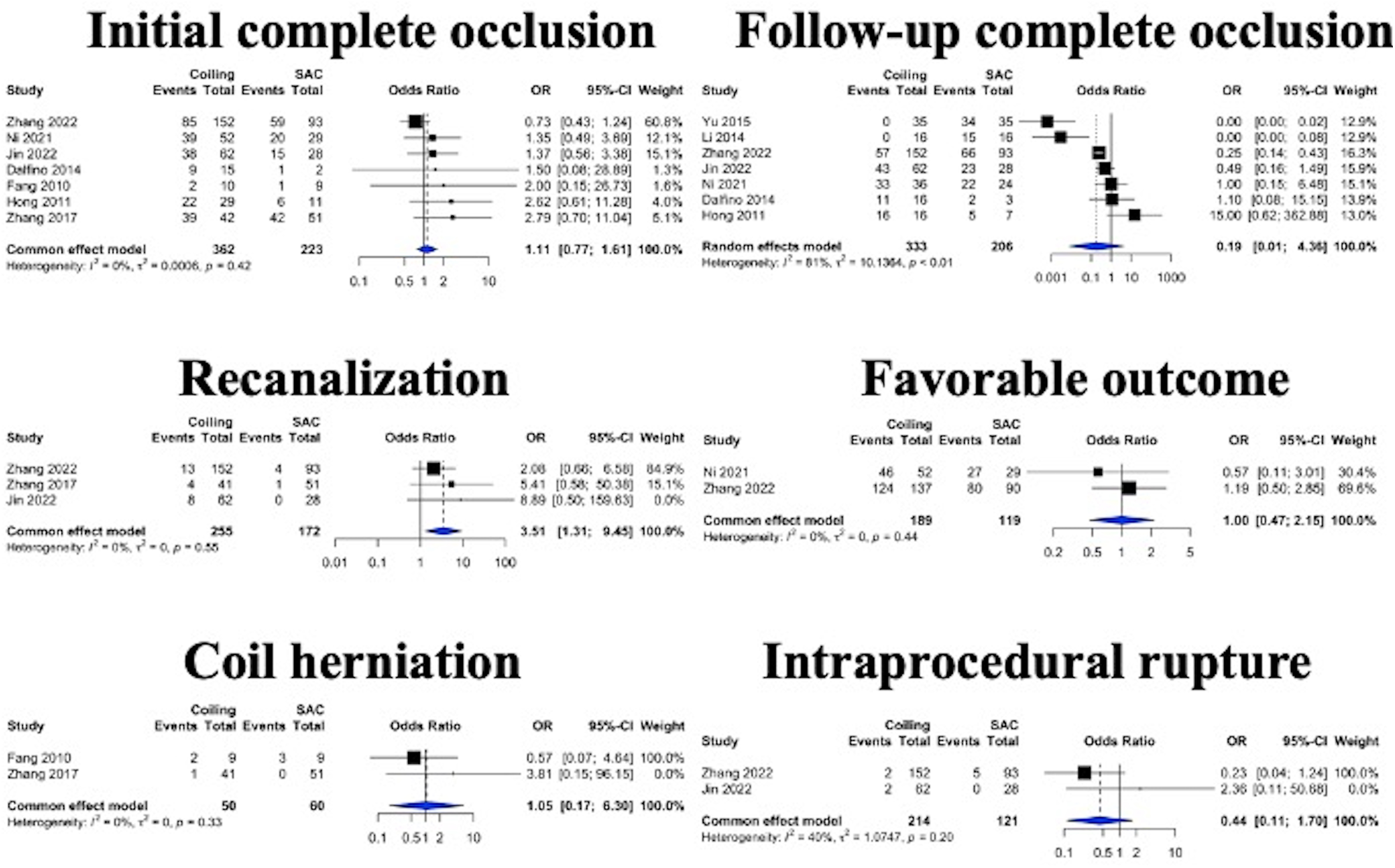
Forest plot for outcomes between coiling and SAC

### Baseline Characteristics

24 included studies with 1355 patients underwent EVT for RVSIAs. The characteristics of the included studies are summarized in Table 1. Thirteen studies were published from China^19, 21, 27–37^; 3 were from USA^38–40^; 2 were from Japan^41, 42^; and 1 each from France^43^, Iran^44^, Korea^45^, Netherland^4^, Taiwan^46^, and UK.^47^ All studies were retrospective cohort except one study. The proportion of male patients ranged from 20% to 65.8%. The mean age ranged from 34.3 to 65.2 years old (standard deviation [SD]:±6.0) and Hunt and Hess grade ranged from 1.4 to 2.8 (SD:±0.35). The median size of aneurysm dome and neck were 2.5 mm (SD:±0.28) and 1.8 mm (SD:±0.27). The mean duration of angiographic follow-up ranged from 4.6 to 28.5 months (standard deviation:±6.7).

**Table 1.**
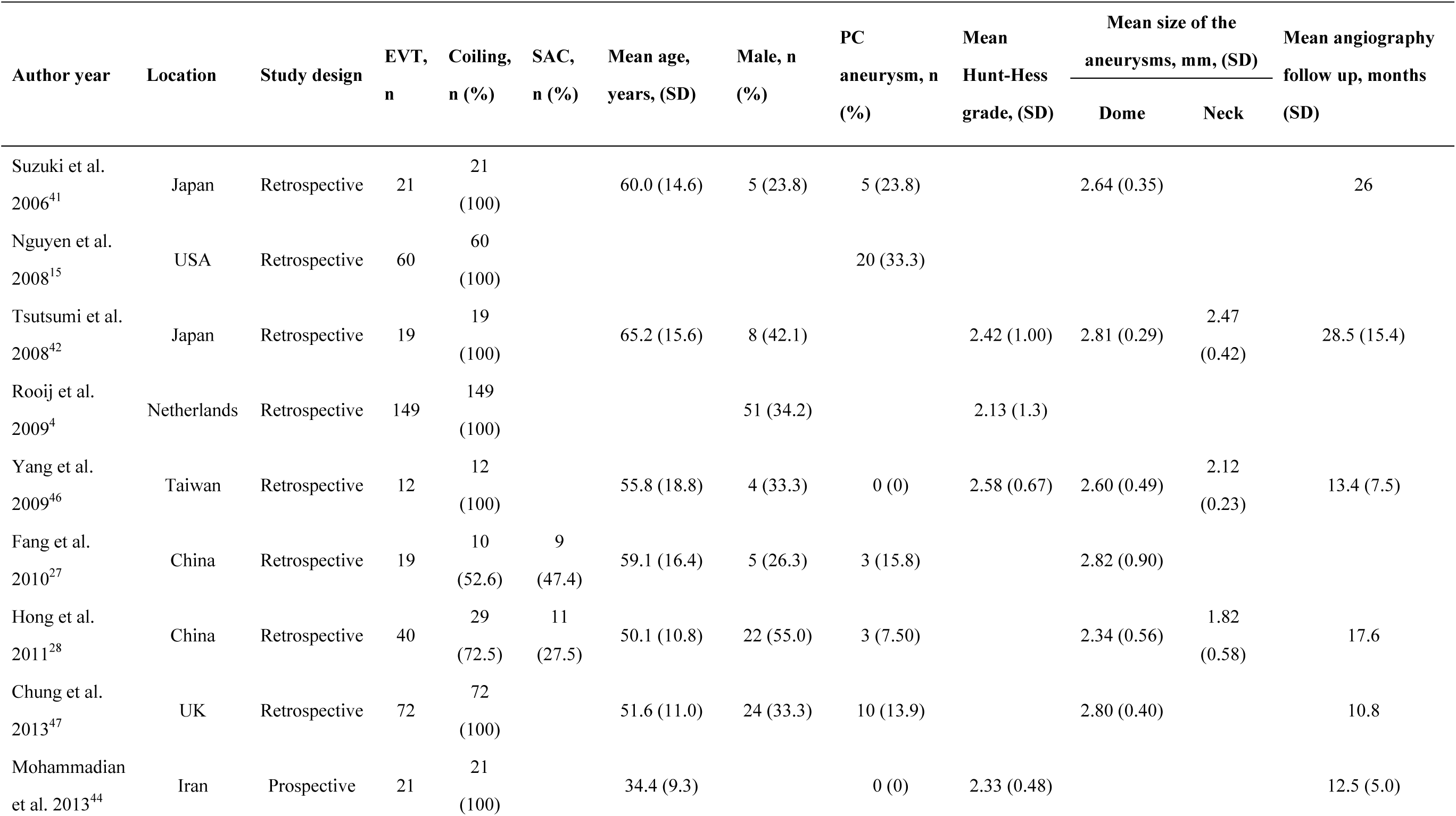
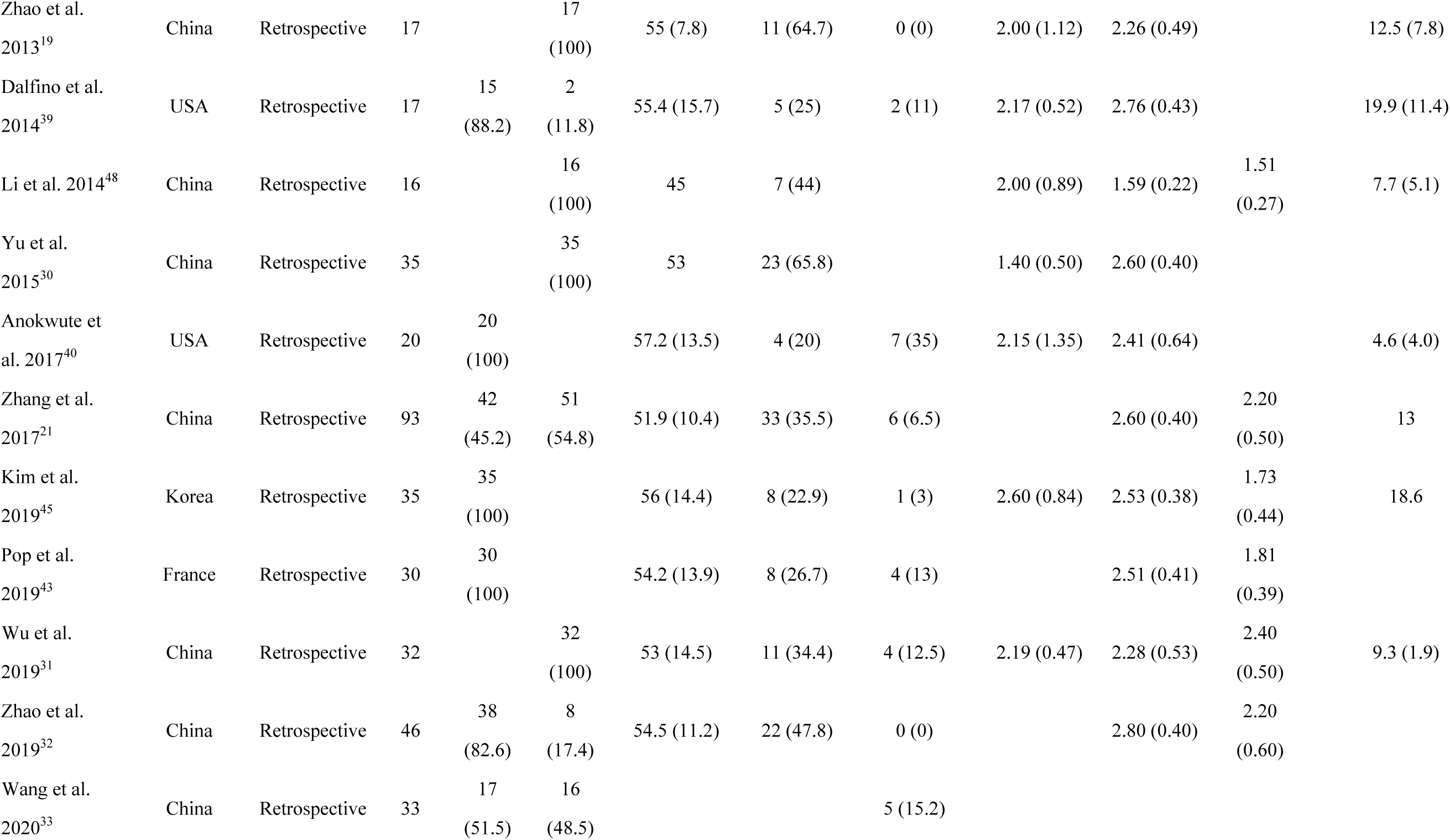
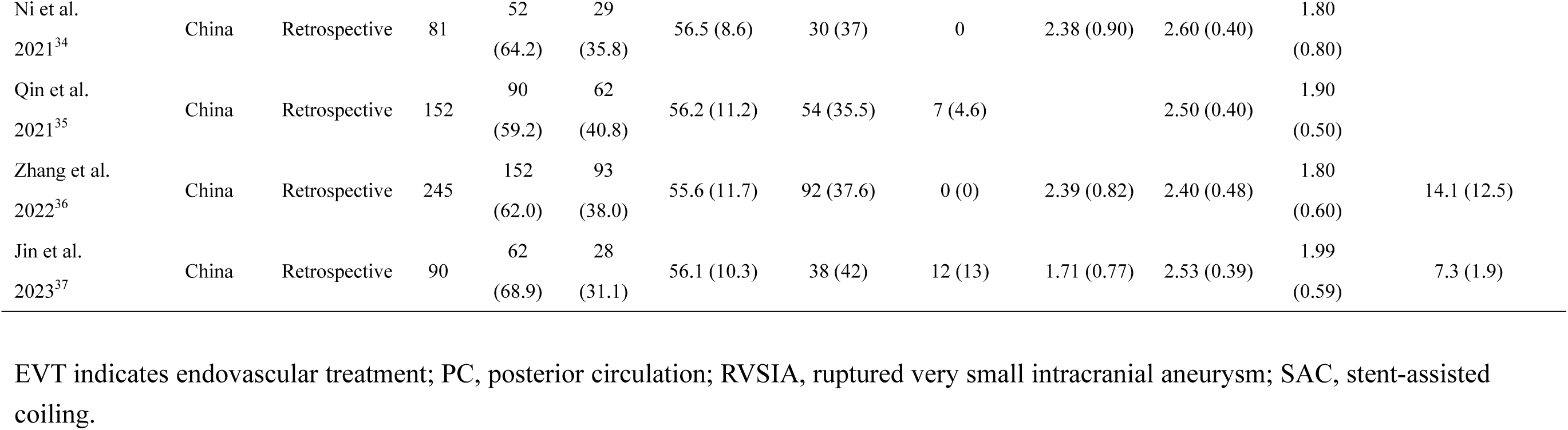
Summary of studies evaluating the EVT of RVSIA

### Risk of Bias

According to the ROBINS quality assessment tool, 10 of our included studies had a moderate risk of bias and the remaining 14 studies had a low risk (Supplement Fig. 1).

## Outcome analysis

### Angiographic and Clinical outcomes

Initial technical failure occurred in 2% (95% CI, 1–5%) of patients. Initial complete aneurysm occlusion rate was 64% (95% CI, 52–74%), while follow-up complete occlusion rate was 85% (95% CI, 74–92%). Recanalization and retreatment were observed in 6% (95% CI, 3–10%) and 3% (95% CI, 2–4%) of the coiled RVSIAs, respectively. Favorable outcome was observed in 91% (95% CI, 89–93%).

### Adverse Events

Overall complications were seen in 9% (95% CI, 7–12%) of subjects. Parent artery vasospasm, coil herniation, and thromboembolism were observed in 6% (95% CI, 5– 9%), 2% (95% CI, 1–8%), and 4% (95% CI, 3–6%), respectively. Intraprocedural re-rupture rate was observed in 4% of RVSIAs (95% CI, 2–7%). Mortality rate was 3% (95% CI, 2–4%).

### Coiling and SAC

Proportion of initial and follow-up complete aneurysm occlusion rates were not significantly different between the coiling and SAC (initial, OR 1.11, [95% CI: 0.77–1.61]; *P*=0.57; follow-up, OR 0.19, [95% CI: 0.01–4.36]; *P*=0.24). Recanalization rate was significantly higher in the coiling group (OR 3.51, [95% CI: 1.31–9.45]; *P*=0.013). Pooling of the 2 studies comparing favorable outcomes between the two groups showed no significant difference (OR 1.00, [95% CI: 0.47-2.15]; *P*=0.99).^34, 36^ Additionally, there were no significance between two groups in procedure-related complications including; coil herniation (OR 1.05: 95% CI: 0.17–6.30, *P*=0.96) and intraprocedural rupture (OR 0.64, [95% CI: 0.11–1.70]; *P*=0.23).

## Discussion

The results of this meta-analysis of the contemporary EVT of RVSIA demonstrate an acceptable low rate of complications, intraprocedural re-ruptures, and retreatment, and high rate of follow-up complete aneurysm occlusion and long-term follow-up favorable outcome. In addition, comparison of coiling and SAC revealed no significant difference in patient outcomes or complications, except for a higher recanalization rate in the coiling group.

Recently, EVT has been widely used in the treatment of ruptured intracranial aneurysms. The findings of the International Subarachnoid Aneurysm Trial (ISAT) suggested that for patients with SAH who were suitable for both EVT and neurosurgical clipping, those with EVT were more likely to survive independently after 1 year than neurosurgical clipping.^49^ On the other hand, the smallest subset of ruptured intracranial aneurysms, those less than 3mm, are generally considered difficult and high-risk to treat with EVT and therefore are often approached with microsurgical clipping. The main challenges of EVT for RVSIA are related to the geometrical challenges they present as they are often broad-necked with a shallow dome. This results in a smaller margin of error in accessing the aneurysm with a microcatheter, as well as difficulty in maintaining microcatheter stability during coil deployment and maintaining the coils within the aneurysm while preventing prolapse into the parent artery. These factors theoretically lead to difficulty in achieving satisfactory coil packing, and higher risk of intraprocedural re-rupture and early rebleeding.^4, 50^ Another previous study showed that in comparison with EVT of larger ruptured aneurysms, tiny aneurysm coiling was related to an increased absolute technical failure rate of 13.7% suggesting that this subset of aneurysms represented the most challenging of all to approach with EVT.^51^ However, as challenging as these aneurysms may be to treat, over time there have been considerable advances in EVT devices and techniques; ranging from improved guide catheters microwires and coils. This contemporary meta-analysis demonstrates high technical success rate (98%) and low intraprocedural rupture rate (4%) among RVSIA treated with EVT. The initial and follow-up complete aneurysm occlusion rate was 64% and 85%. During follow-up, recanalization observed in only 6% and retreatment was performed in just 3%. Favorable outcome was achieved in 91% of subjects and mortality rate was 3%. In addition to re-rupture, other intraprocedural events such as thromboembolism, coil herniation, and parent artery vasospasm are also significant issues to be avoided. In this analysis, the incidence of thromboembolism, coil herniation, and parent artery vasospasm was 4%, 2%, and 6%, respectively. Compared to the results of previous meta-analysis, the rate of initial complete occlusion, intraprocedural rupture and retreatment was lower, the rate of long-term favorable outcome was higher, and follow-up complete occlusion rate was similar to our meta-analysis (Table 2). This could be in part due to a more conservative coiling approach by treating physicians attempting to avoid the most dreaded complication of all intraprocedural re-rupture.

**Table 2.**
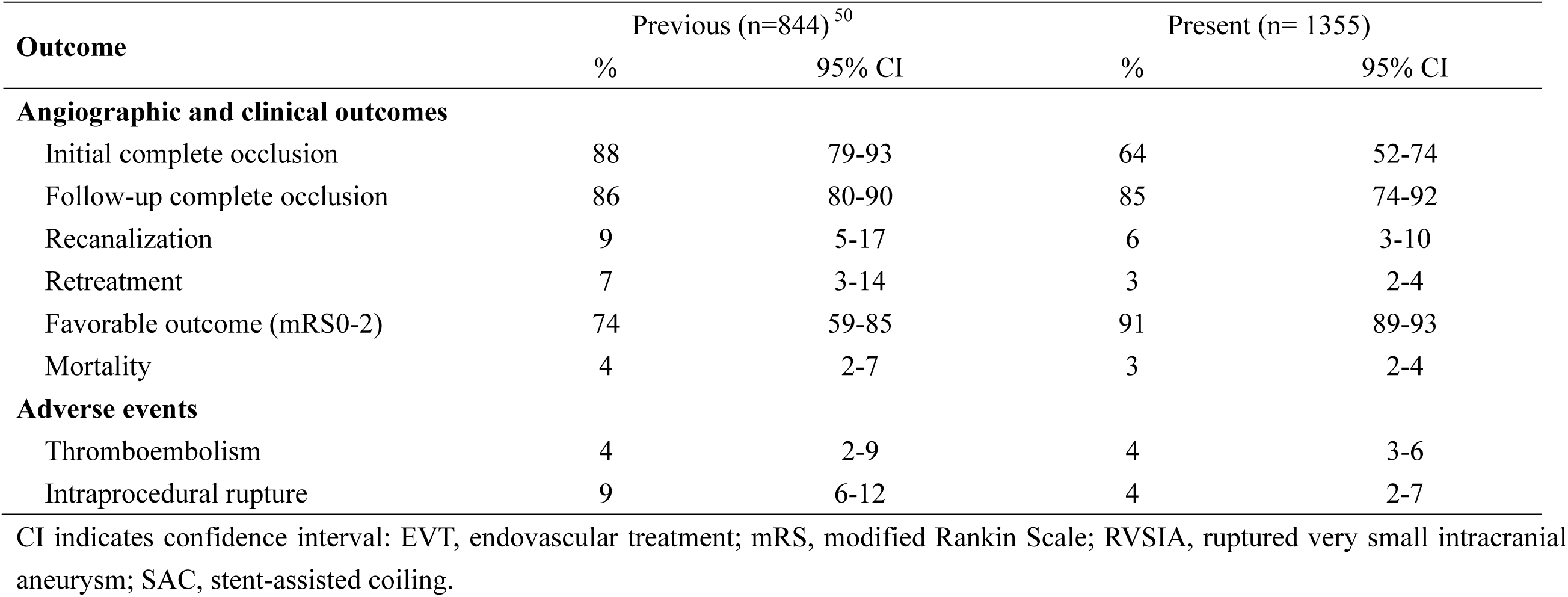
Comparison of the results of previous and present study about EVT of RVSIAs

The efficacy and safety of EVT of RVSIAs may be due to not only the accumulated experience of neurointerventional physicians but also the introduction of new microcatheters and microguidewires with improved trackability, pushability, torque, and novel hydrophilic coatings, which could help neurointerventionalists safely access a very small aneurysm dome. Furthermore, the evolution of various kinds of soft and extra soft coils could also have contributed to the higher success rate of EVT of RVSIA. While relatively safe and effective, 9% and 4% frequency of overall complications and retreatment are acceptable but not insignificant, therefore, neurointerventionalists should keep continue to improve clinical and radiological outcomes in the management of RVSIA with EVT.

Stent placement could help maintain coils inside the aneurysm, thus increasing the coil packing density and decreasing the stent porosity at the neck plane. Stent struts across the aneurysm neck also could provide the structural basis for endothelialization and moderate the hemodynamic condition.^21^ Since the majority of RVSIAs are wide-necked, SAC is often needed from a purely mechanical and geometric sense. However, in the acute stage of subarachnoid hemorrhage, the patient is in a hypercoagulable state and, therefore, SAC may increase the risk of thromboembolic events as well as hemorrhagic complications such as external ventricular drain site bleeds. In fact, Roh et al. reported that the rate of thromboembolic complications in stent-assisted coil embolization of ruptured intracranial aneurysms was approximately twice as high as that in non-stent coil embolization.^52^ This meta-analysis showed no significant difference between coiling and SAC of RVSIAs in the rate of initial and follow-up complete aneurysm occlusion, intraprocedural rupture, coil herniation, and favorable outcome.

Although both coiling and SAC could be selected in the treatment of RVSIAs, physician should recognize that while recanalization rate might be significantly higher in the coiling alone group, this potential benefit must be weighed against the risk of exposing the patient to dual antiplatelet therapy in the acute ruptured setting.

### Limitations

This meta-analysis has several limitations. First, moderate heterogeneity was found. Second, there is also bias inherent in the retrospective nature of most studies included. Third, another periprocedural complications, such as extraventricular drainage tract hemorrhage, intraventricular hemorrhage, and aSAH-induced delayed cerebral vasospasm, and antiplatelet managements are also important issues in the management of aSAH, though, these variables could not be sufficiently investigated due to lack of data. Fourth, whether balloon-assisted coiling was performed could not be collected due to missing data. In this regard, some studies showed that balloon use was related to thromboembolic complications and intraprocedural rupture^4, 41^, though the other indicated that it was not associated with intraprocedural rupture.^53^ Fifth, thorough investigation of delayed post-treatment aneurysm rupture, a relevant concern, was not possible, due to lack of long term data. Finally, mortality, retreatment, thromboembolism, parent artery vasospasm could not be compared between coiling and SAC due to the lack of data. Nevertheless, this contemporary meta-analysis is currently the largest study assessing the EVT of RVSIAs with more than 1300 subjects.

## Conclusion

This meta-analysis demonstrates that EVT is a feasible, effective, and safe approach for the treatment of RVSIA and is associated with favorable clinical outcomes in both the short and long term. Further well-designed randomized controlled trials with large sample size are warranted to verify our findings.

## Data Availability

The data that support the findings of this study are available from the corresponding author, Alejandro M Spiotta, upon reasonable request.

## Acknowledgement

NA

## Abbreviations

EVT: endovascular treatment
RVSIA: ruptured very small intracranial aneurysm
SAC: stent-assisted coiling

